# Genomic and phenotypic characterization of Multisystem Inflammatory Syndrome in Children (MIS-C): a prospective multicenter study from the Middle East

**DOI:** 10.1101/2021.11.30.21267109

**Authors:** Walid Abuhammour, Lemis Yavuz, Ruchi Jain, Khawla Abu Hammour, Ghalia F. Al-Hammouri, Maha El Naofal, Nour Halabi, Sawsan Yaslam, Sathishkumar Ramaswamy, Alan Taylor, Deena Wafadari, Ali Alsarhan, Hamda Khansaheb, Zulfa Omar Deesi, Rupa Murthy Varghese, Mohammed Uddin, Hanan Al Suwaidi, Abdulmajeed Alkhaja, Laila Mohamed AlDabal, Tom Loney, Norbert Nowotny, Abdulla Al Khayat, Alawi Alsheikh-Ali, Ahmad Abou Tayoun

## Abstract

**Importance:** Clinical, genetic and laboratory characteristics of patients with multisystem inflammatory syndrome in children (MIS-C) in the Middle East have not yet been documented.

**Objective:** To uncover rare genetic variants contributing to MIS-C in patients of primarily Arab and Asian origin.

**Design, Setting, and Participants:** A prospective multicenter cohort study was conducted between September 2020 and August 2021 in the United Arab Emirates and Jordan. Forty-five patients meeting the case definition of MIS-C, and a matched control group of twenty-five healthy children with a confirmed severe acute respiratory syndrome coronavirus 2 (SARS-CoV-2) infection status, were recruited. Whole Exome Sequencing (WES) in all 70 participants was performed to identify rare likely deleterious variants in patients with MIS-C and to correlate genetic findings with the clinical course of illness.

**Exposures:** SARS-CoV-2

**Main Outcomes and Measures:** Fever, organ system complications, laboratory biomarkers, WES findings, treatments, and clinical outcomes. Mann–Whitney *U* test was used to assess the association between genetic variations and MIS-C attributes. Fisher’s exact test was used to compute the genetic burden in MIS-C relative to controls.

**Results:** In 45 MIS-C patients (23 boys [51.1%]; average age, 7 years [range, 2-14 years]), key inflammatory markers were significantly dysregulated in all patients. Mucocutaneous and gastrointestinal manifestations were reported in 80% (95% CI 66% to 89%) while cardiac and neurological findings were reported in 49% (95% CI 35% to 63%) and 31% (95% CI 19.5% to 45.6%) of patients, respectively. Rare, likely deleterious heterozygous variants in immune-related genes including *TLR3, TLR6, IFNAR2, IL22RA2, IFNB1*, and *IFNA6*, were identified in 19 patients (42.2%, 95% CI 29% to 56.7%), of whom seven had more than one variant. There was significant enrichment of genetic variants in patients relative to the control group (29 versus 3, P<.0001). Those variants were significantly associated with earlier onset of disease (31.5%, 95% CI 15.4% to 54% of patients with versus 7.7%, 95% CI 2% to 24% without genetic findings were < 3 years) and resistance to treatment (42%, 95% CI 23% to 64% of patients with versus 11.5%, 95% CI 4% to 29% of patients without genetic findings received two doses of IVIG).

**Conclusions and Relevance:** Rare, likely deleterious genetic variants contribute to MIS-C onset and management.

**Key Points:** *Question:* What are the clinical, genetic, and laboratory characteristics of multisystem inflammatory syndrome in children (MIS-C) of the Middle East?

*Findings:* In this prospective study of 45 MIS-C patients of primarily Arab and Asian origins, we show that the clinical course was consistent with that of previously characterized patients from other backgrounds. However, we find an enrichment of rare, likely deleterious immune-related genetic variants, in MIS-C patients, and an association of genetic status with MIS-C onset and resistance to treatment.

*Meaning:* Comprehensive genetic profiling of MIS-C patients of diverse ancestries is essential to characterize the genetic contribution to this disease.

## Introduction

Multisystem Inflammatory Syndrome in Children (MIS-C) is a critical and potentially life-threatening complication of COVID-19 in pediatric settings. MIS-C was first identified in Europe, initially termed as SARS-CoV2-related pediatric multisystem inflammatory syndrome, and later referred to as MIS-C by the United States Centers for Disease Control and Prevention (CDC) (1). Studies revealed that 70% of all children who were diagnosed with MIS-C were positive for SARS-CoV-2 infection by RT-PCR, antibody testing, or both (2, 3, 4, 5). The majority of patients had fever for more than 4 days (2, 6, 7, 8), and the most common presenting features included gastrointestinal symptoms (80-100%) (7, 9, 10, 8), mainly vomiting, abdominal pain, and/or diarrhea. In addition, mucocutaneous manifestations such as conjunctivitis and rash with neurological findings were present (2, 9, 3, 7, 8). In 2020, the estimated incidence of laboratory-confirmed SARS-CoV-2 infection in individuals <21 years old was 322 per 100,000 and the incidence of MIS-C was 2 per 100,000 (6).

As presented by Riphagen et al., hyperinflammatory shock is a common element in MIS-C (11). More than 60% of the patients were managed therapeutically with intravenous immunoglobulin (IVIG) (12). Steroids were used in all studies ranging from 10-95% of the included patients. On the other hand, aspirin was used in few studies (2,11,13). Most patients responded favorably to therapeutic management with an improvement in vital signs and cardiac dysfunction.

In a recent study, exome sequencing of two MIS-C patients delineated a role of one gene, *SOCS1*, in the IFN pathway, in predisposing individuals to infection-associated autoimmune cytopenias (14). Similarly, another exome sequencing study identified defects in X-linked inhibitor of apoptosis, *XIAP*, and in *CYBB*, encoding cytochrome b-245, in 3 out of 18 sequenced MIS-C patients (15), whereas RNA sequencing of MIS-C patients revealed dysregulated cytotoxic lymphocyte responses to SARS-Cov-2 infection as major drivers (16). These studies suggest dysregulation of the inflammatory response as a characteristic of MIS-C, though they were limited to a small number of patients who were most often of European origin.

Here, we performed whole exome sequencing in 45 patients with MIS-C and 25 healthy children who were infected with SARS-CoV-2 but remained asymptomatic or developed only mild clinical symptoms (control group). We focused on rare deleterious variants in IFN/cytokines/immune related genes between MIS-C patients and the control group and assessed any associations between genetic variants and MIS-C disease onset, symptoms, laboratory markers, treatments, and clinical outcomes (**Figure 1**).

**Figure 1.**
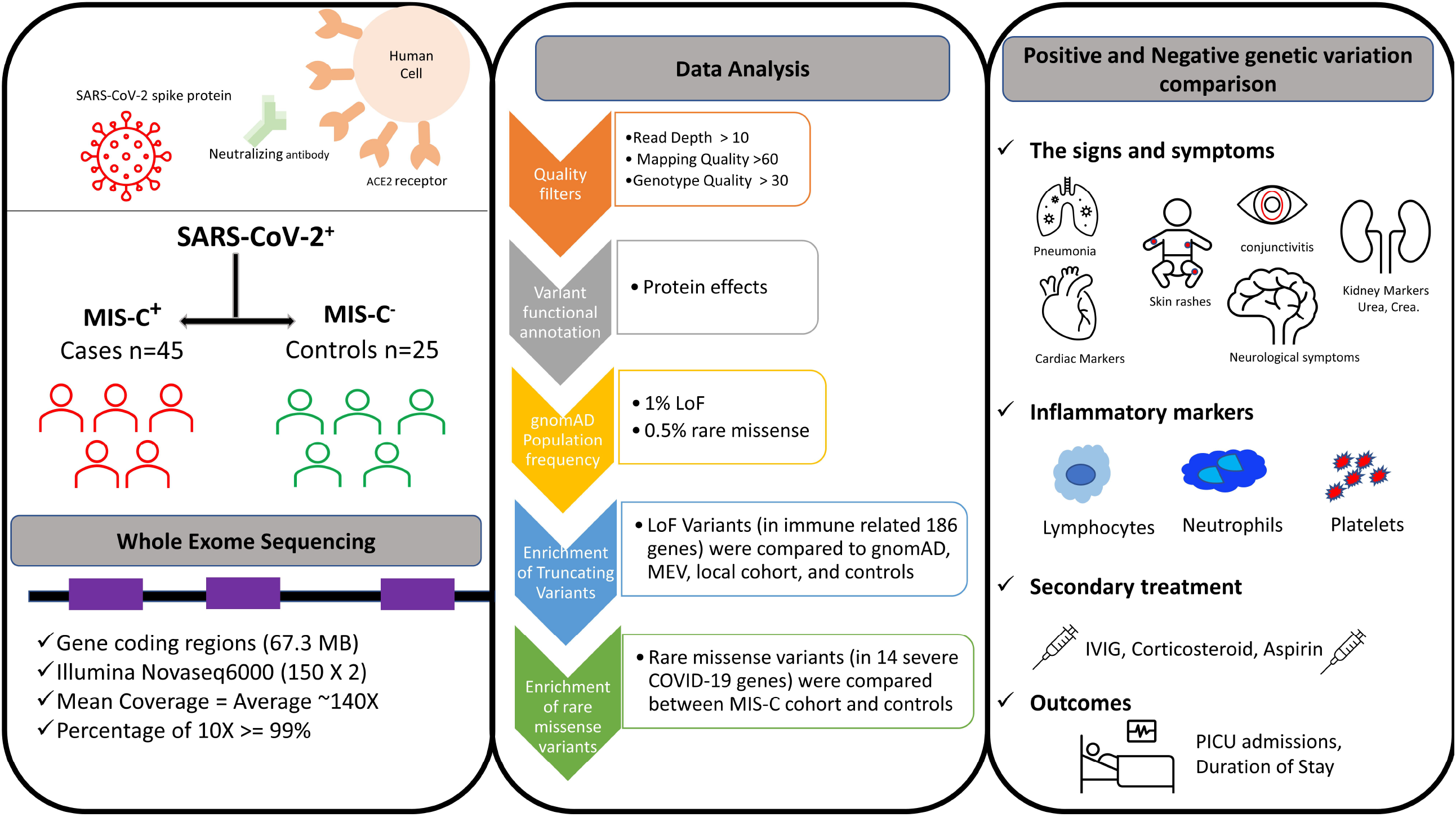
Graphical representation of study design, participants, sequencing protocol, bioinformatic analysis, and genomic and clinical characterization of patients with MIS-C. SARS-CoV-2; severe acute respiratory syndrome coronavirus 2; ACE2, angiotensin converting enzyme 2; MIS-C, multisystem inflammatory syndrome in children; LoF, Loss of function; gnomAD, Genome Aggregation Database; MEV, Middle East Variome database; IVIG, Intravenous immunoglobulin; PICU, Pediatric intensive care unit.

## Subjects and Methods

### Patients

This study was approved by the Dubai Scientific Research Ethics Committee—Dubai Health Authority (Approval number DSREC-07/2020_04) and the Institutional Review Board of the Specialty Hospital, Jordan (Approval Number 5/1/T/104046). Patients (and their guardians) recruited in Dubai or Jordan provided written consent for their de-identified data to be used for research, and this study was performed in accordance with the relevant laws and regulations that govern research in both countries.

We recruited 70 patients, all children between 1 day to 18 years old, divided into two groups for this study. First group included 45 patients diagnosed with MIS-C. Inclusion criteria for this group were consistent with the case definition of Multi-System Inflammatory Syndrome in Children set by the World Health Organization (WHO) or CDC (**Supplementary Table 1**). All patients had evidence of SARS-CoV-2 infection either by RT-PCR, SARS-CoV-2 antibodies, or recent exposure to a confirmed case of COVID-19. At least two organs were affected by the illness, and their blood profile revealed increased inflammatory markers. Exclusion criteria were applied to all patients with another diagnosis that can affect their course, such as congenital heart disease, failure to thrive, or other syndromes. The second group included 25 healthy children (aged <8 years), hereafter referred as Controls, were recruited at same time, had SARS-CoV-2 infection which was confirmed by RT-PCR yet were asymptomatic or experienced mild symptoms.

### Clinical and Demographic Information

De-identified patient information were extracted from electronic medical records at Al Jalila Children’s Hospital (Dubai, UAE), Latifah Hospital (Dubai, UAE) and The Specialty Hospital (Jordan) where those patients were treated and recruited for the study. Variables included demographic information, signs and symptoms on admission, inflammatory markers with cytokine profile, cardiac manifestation, course of illness, admission to the Paediatric Intensive Care Unit (PICU), treatment, and outcome.

### Whole Exome Sequencing

Whole Exome Sequencing (WES) was performed in the genomic laboratory at Al Jalila Children’s Hospital. DNA was extracted from peripheral blood cells using standard DNA extraction protocols (Qiagen, Germany). Following fragmentation by ultra-sonication (Covaris, USA), genomic DNA was processed to generate sequencing-ready libraries of short fragments (300-400bp) using the SureSelect^XT^ kit (Agilent, USA). RNA baits targeting all coding regions were used to enrich for whole exome regions using the SureSelect Clinical Research Exome V2 kit (Agilent, USA). The enriched libraries underwent next generation sequencing (2 × 150bp) using the SP flow cell and the NovaSeq platform (Illumina, USA).

### Bioinformatics Analysis and Variant Filtration

Sequencing data was processed using an in-house custom-made bioinformatics pipeline to retain high quality sequencing reads with greater then 10X coverage across all coding regions. High quality variants with Read Depth > 10, Genotypic Quality > 30, Mapping Quality > 60 were retained for downstream analysis (17). We prioritized rare variants in 182 signature genes that may possibly be implicated in disease progression of MIS-C, mainly genes that are associated with immune responses, including cellular response to cytokine, cell mediation of immunity, immune and interferon signaling pathways from reported literature (18, 19, 20, 21). We used three filters to retain: a) truncating or loss of function (LoF) variants in the 182 inflammatory/immune related genes with deleterious effect on RefSeq canonical transcripts, and allele frequency <1% in the Genome Aggregation Database (gnomAD); b) homozygous missense or loss of function variants across the 186 genes, and allele frequency <1% in gnomAD; and c) missense variants in a subset of genes (N =14) recently associated with severe COVID-19 (18, 19) and gnomAD allele frequency <0.5%. We applied similar filters to WES data from controls (N = 25) and compared these variants between patients with MIS-C and controls.

### Enrichment Analysis

Enrichment of rare, likely deleterious genetic variants in patients with MIS-C was determined by comparing the proportion of functional alleles (nonsense, frameshift, missense, and canonical ±1,2 splicing) in the MIS-C group to that in controls, and *p*-values were calculated using the Fisher’s exact test (GraphPad Prism v9.2.0). Similarly, the number of patients with more than one heterozygous variant were compared between the MIS-C and control groups, as another measure of the burden of genetic variation in patients.

Furthermore, to rule out the possibility that the genes affected with truncating or LoF variants in patients with MIS-C were simply tolerant to such variation, we quantified the fractions of such variants in those genes in general populations, namely gnomAD (22), MEV (Middle East Variome database, created inhouse by assembling sequencing data from Qatar (23) and GME, Greater Middle East) (24), an internal local cohort (UAE healthy parents exome sequencing data) and controls, and compared to that in MIS-C. For each gene, total allele count was divided by maximum allele number in the database and Fisher’s exact test was performed using Graph Pad Prism v9.2.0.

### Statistical analysis

We divided patients into two groups “positive” and “negative” based on presence or absence of genetic variation, respectively, and performed Mann Whitney *U*-test to compare clinical and pathological findings (mucocutaneous, gastrointestinal and neurological), inflammatory markers (WBC, Hb, CRP, AST, ALT, Neutrophils, Lymphocytes, Platelets, D-Dimer, ESR, Albumin, Ferritin, Fibrinogen, Urea, Creatinine), treatment, management and outcomes (Admission to PICU, IVIG doses etc.) using Graph Pad Prism v9.2.0. We did not impute missing data. Categorical variables were compared using the Fisher’s exact test. Tests were two-tailed, and *p*-value <0.05 was considered significant.

### Pathway and Protein-protein interaction Analysis

String (25) was used for pathway enrichment and protein-protein interaction (PPI) network analysis. The network nodes correspond to proteins while edges represent the functional associations between nodes based on curated databases, experimental data, or predicted interactions based on co-expression data and protein homology.

## Results

### Demographic and clinical characteristics of patients with MIS-C

Forty-five children who met the case definition of MIS-C (**Supplementary Table 1**) were prospectively recruited into the cohort. Males and females were equally represented (49% females), with an average age of 7 years (range 2 – 14 years) (**Figure 2A**). Most patients had Middle Eastern origin (66.67%). More than half (55.5%) were Arab-descendants, mainly Emiratis, Jordanians, or Egyptians, and 20% were Asians (**Figure 2B**).

**Figure 2.**
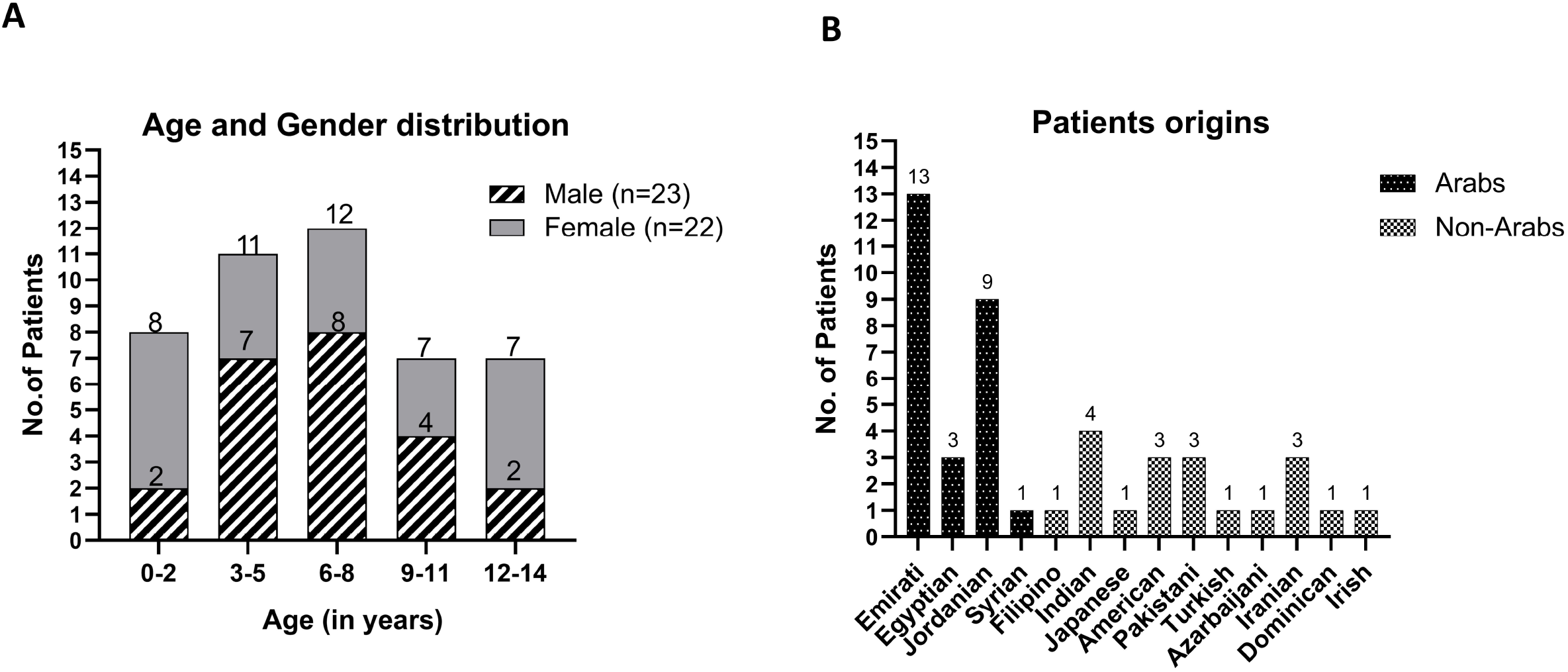
Age, gender and origin of MIS-C patients. **A**, Age and gender distribution of MIS-C patients, x-axis represents age (in years), and y-axis represents number of patients. Grey filled areas represent females while dashed areas represent males. **B**, Origin of patients with MIS-C, x-axis shows country of origin while y-axis shows number of patients. Black filled bars represent Arabs while grey filled bars represent non-Arabs.

Thirty-six (80%) patients had evidence of SARS-CoV-2 infection either by PCR or antibody, while nine (20%) had exposure to the virus through contact with COVID-19 patients. Average duration of fever was around 5 days (**Table 1** and **Supplementary Table 2**) and all patients had laboratory evidence of inflammation including lymphopenia and/or significantly elevated inflammatory markers, such as CRP, ESR, D-dimer, ferritin, fibrinogen, and IL-6 (**Table 2**).

**Table 1:** Clinical characteristics of MIS-C cohort (N=45)

|  |  |
| --- | --- |
| Average age in years (range) | 7 (2-14) |
| Fever (%) | 100% |
| Evidence of SARS-CoV-2 (PCR, antibodies, exposure) (%) | 100% |
| Raised inflammatory markers (CRP, PCT, ESR) (%) | 100% |
| Rash or bilateral non-purulent conjunctivitis or muco-cutaneous inflammatory signs (% , N) | 80%, 36 |
| Shock (% , N) | 42%, 19 |
| Myocardial dysfunction, pericarditis, valvulitis, or coronary abnormalities (including ECHO findings or elevated Troponin/NT-proBNP) (% , N) | 49%, 22 |
| Acute gastrointestinal problems (% , N) | 80%, 36 |
| Neurological symptoms (% , N) | 31%, 14 |
| Respiratory symptoms (% , N) | 26.6%, 12 |
| Kidney Markers (% , N) | 42%, 19 |
| Abbreviations: Acute gastrointestinal problems (vomiting, abdominal pain, diarrhea). Neurological symptoms (headache, blurred vision, Papilledema, meningitis, MRI abnormalities). Respiratory symptoms (pneumonia or pleural effusion). Kidney Markers (US abdomen) |  |

**Table 2.**
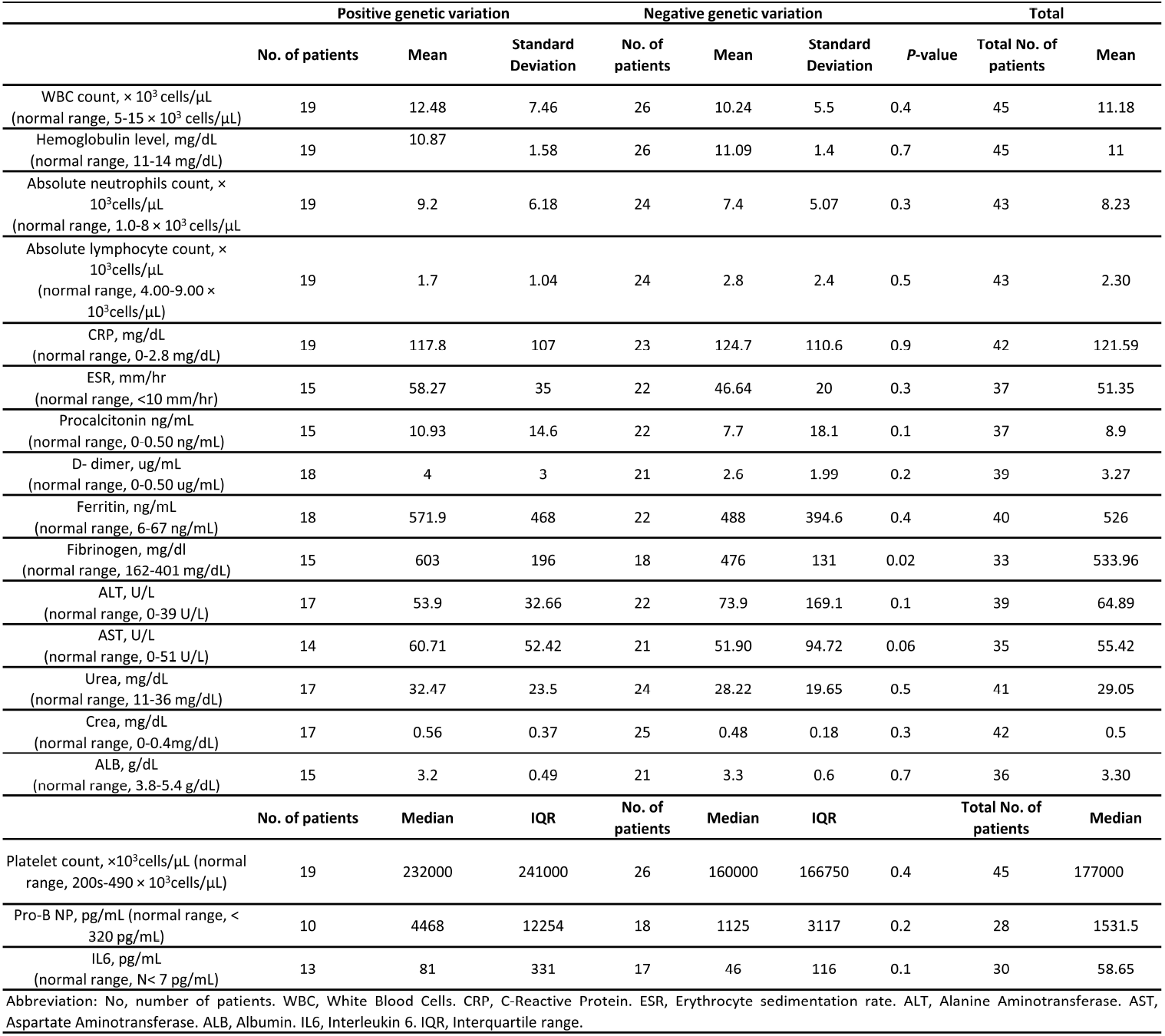
Inflammatory Markers in MIS-C Patients on Admission.

Gastrointestinal symptoms, mainly vomiting, abdominal pain and/or diarrhoea, and mucocutaneous involvement, characterized by skin rash, inflammation of oral mucosa, conjunctivitis, and/or extremity findings including edema of hands and feet, were the most common clinical findings, each was reported in 36 (80%) patients. Neurological symptoms, defined by headache, blurred vision, papilledema, meningitis, and/or MRI abnormalities, were reported in 14 (31%) patients while respiratory symptoms, including pneumonia with or without pleural effusion were reported in 12 (26.6%) patients (**Table 1**).

Cardiac involvement was reported in 22 patients (49%), while nineteen (42%) patients had hypotension and were in shock (**Table 1**). The clinical manifestations in our cohort were consistent with findings in the scientific literature (26, 27).

### Genetic and enrichment analysis

Whole exome sequencing was performed on 45 patients with MIS-C and 25 controls. All patients had average coverage >100X with >99% of coding regions covered with at least 10 reads. Three filter sets were used to retain rare, truncating, or homozygous variants in a broad set of 186 immune-related genes (**Supplementary Table 4**), and rare, missense variants in a subset of genes (N = 14) recently shown to be associated with severe COVID-19 (**Figure 1** and Methods).

We detected twenty truncating heterozygous variants affecting sixteen immune-related genes (*CD84, CD163, IFI44, IFI44L, IFIH1, IFNA21, IFNA4, IFNA6, IFNB1, IL22RA2, IRAK3, LY9, NLRP12, NLRP2, RAB27A*, and *TLR6*) in fourteen patients with MIS-C (**Table 3**). Those genes were not similarly enriched for truncating variants in the general population like in gnomAD, in a combined set of exome sequencing data from the Greater Middle East (GME) variome project (N = 1111) (24) and a Qatari cohort (N = 1005) (23), or in our internal cohort of healthy individuals (N =124) (**Figure 3A**). Finally, using a similar filtration strategy in the control group, we detected only three truncating variants across all 186 genes (versus 20 in MIS-C patients) (P<0.05, Fisher’s exact test) further confirming the significant burden of such variants in the MIS-C group (**Figure 3A** and **B**). Furthermore, six of the fourteen MIS-C patients with truncating variants harbored two such variants in two different genes (**Table 3**), including two patients (COVGEN-7 and COVGEN-36) who were double heterozygotes for truncating variants in *IL22RA2* and *IFI44L*, and another patient (COVGEN-30) with two heterozygous variants in *IFNA6* and *IFNA21*.

**Table 3:** Genetic Findings in MIS-C cohort.

| Table 3: Genetic Findings in MIS-C cohort |  |  |  |  |  |  |  |  |
| --- | --- | --- | --- | --- | --- | --- | --- | --- |
| Case ID | Chromosome coordinates | Gene(s) | Transcript | cDNA | Protein Effect | Zygosity | Effect | Disease Pathway |
| COVGEN-27 | chr19:54301595 | *NLRP12* | NM_144687.3 | c.2828_2829dupTC | p.Arg944Serfs*6 | Het | Frameshift | NOD-like receptor signaling pathway |
|  | chr4:38830758 | *TLR6* | NM_006068.4 | c.337C>T | p.Gln113Ter | Het | Nonsense | Toll-like Receptor Signaling Pathway |
|  | chr19:50163998 | *IRF3* | NM_001571.6 | c.1070C>T | p.Pro357Leu | Het | Missense | Toll-like Receptor Signaling Pathway |
| COVGEN-18 | chr12:66641621 | *IRAK3* | NM_007199.3 | c.1461delC | p.Asn487Lysfs*10 | Het | Frameshift | IL-1 signaling pathway |
|  | chr1:160784531 | *LY9* | NM_002348.4 | c.1052_1053delAT | p.His351Argfs*22 | Het | Frameshift | Interleukin-2 signaling pathway |
|  | chr4:187003729 | *TLR3* | NM_003265.3 | c.889C>G | p.Leu297Val | Het | Missense | Toll-like Receptor Signaling Pathway |
|  | chr14:103369742 | *TRAF3* | NM_003300.4 | c.1111G>A | p.Ala371Thr | Het | Missense | Toll-like Receptor Signaling Pathway |
| COVGEN-36 | chr6:137479566 | *IL22RA2* | NM_052962.3 | c.115C>T | p.Arg39Ter | Het | Nonsense | Cytokine Signaling in Immune system |
|  |  | *IFI44L* | NM_006820.4 | c.873T>A | p.Tyr291Ter | Het | Nonsense | Immune response |
| COVGEN-7 | chr6:137479566 | *IL22RA2* | NM_052962.3 | c.115C>T | p.Arg39Ter | Het | Nonsense | Cytokine Signaling in Immune system |
|  |  | *IFI44L* | NM_006820.4 | c.873T>A | p.Tyr291Ter | Het | Nonsense | Immune response |
| COVGEN-6 | chr2:163133953 | *IFIH1* | NM_022168.4 | c.2016delA | p.Asp673Ilefs*5 | Het | Frameshift | Induction of type I interferons and proinflammatory cytokines |
| COVGEN-23 | chr19:54299165 | *NLRP12* | NM_144687.3 | c.3046C>T | p.Arg1016* | Het | Nonsense | NOD-like receptor signaling pathway |
| COVGEN-29 | chr9:21077466 | *IFNB1* | NM_002176.4 | c.403G>T | p.Gly135* | Het | Nonsense | Toll-like Receptor Signaling Pathway |
| COVGEN-30 | chr9:21350700 | *IFNA6* | NM_021002.2 | c.187C>T | p.Glu126Ter | Het | Nonsense | Toll-like Receptor Signaling Pathway |
|  | chr9:21166236 | *IFNA21* | NM_002175.2 | c.376G>T | p.Gln63Ter | Het | Nonsense | Toll-like Receptor Signaling Pathway |
| COVGEN-8 | chr2:163136505 | *IFIH1* | NM_022168.4 | c.1641+1G>C | p.? | Het | - | Induction of type I interferons and proinflammatory cytokines |
| COVGEN-13 | chr1:160535263 | *CD84* | NM_003874.4 | c.319delT | p.Tyr107Thrfs*5 | Het | Frameshift | Regulation of innate and adaptive immune response |
|  | chr12:7635263 | *CD163* | NM_203416.4 | c.3223C>T | p.Arg1075Ter | Het | Nonsense | Anti-inflammatory role |
| COVGEN-16 | chr19:55493698 | *NLRP2* | NM_017852.5 | c.632dupT | p.Tyr212Valfs*57 | Het | Frameshift | Activation of proinflammatory caspases |
|  | chr1:79101171 | *TRIM69* | NM_182985.5 | c.1404C>A | p.Phe468Leu | Het | Missense | Antigen processing and presentation |
| COVGEN-38 | chr15:55516202 | *RAB27A* | NM_004580.5 | c.352C>T | p.Gln118Ter | Het | Nonsense | Immune responses |
| COVGEN-39 | chr1:79121088 | *IFI44* | NM_006417.5 | c.732C>G | p.Tyr244Ter | Het | Nonsense | Immune responses |
| COVGEN-45 | Chr9:21187471 | *IFNA4* | NM_021068.3 | c.60T>A | p.Cys20* | Het | Nonsense | Toll-like Receptor Signaling Pathway |
| COVGEN-33 | chr4:187003729 | *TLR3* | NM_003265.3 | c.889C>G | p.Leu297Val | Het | Missense | Toll-like Receptor Signaling Pathway |
| COVGEN-25 | chr15:45059871 | *TRIM69* | NM_182985.5 | c.1404C>A | p.(Phe468Leu) | Het | Missense | Antigen processing and presentation |
| COVGEN-20 | chr21:34632925 | *IFNAR2* | NM_001289125.3 | c.733G>C | p.Gly245Arg | Het | Missense | Toll-like Receptor Signaling Pathway |
| COVGEN-17 | chr21:34632925 | *IFNAR2* | NM_001289125.3 | c.733G>C | p.Gly245Arg | Het | Missense | Toll-like Receptor Signaling Pathway |
| COVGEN-43 | chr21:34635648 | *IFNAR2* | NM_001289125.3 | c.1391A>C | p.Asn464Thr | Het | Missense | Toll-like Receptor Signaling Pathway |

**Figure 3.**
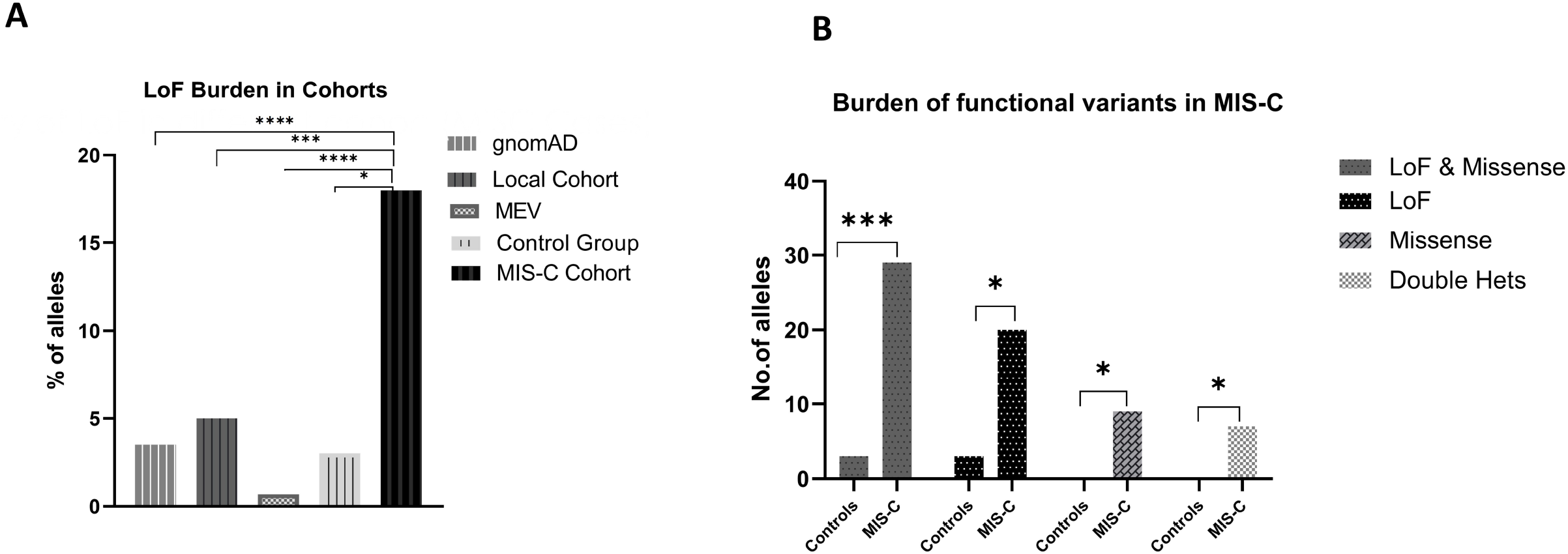
Burden of immune-related LoF and missense variants in MIS-C patients. **A**, Frequency of truncating variants in the 16 genes identified in MIS-C cohort relative to different populations, y axis representing % of alleles. **B**, Number of LoF variants, Missense variants, or both, and number of double heterozygotes in the control group and MIS-C patients, y-axis represents number of alleles. \**P* < .05; \*\**P* < .01; \*\*\**P* < .001; \*\*\*\**P* < .0001by the Mann-Whitney test.

We also identified six rare missense variants in nine patients with MIS-C (three of whom also carried truncating variants from above analysis) in five of the fourteen genes (*IFNAR2, IRF3, TLR3, TRAF3*, and *TRIM69*) known to cause severe COVID-19 (**Table 3**), and none in the control group. Interestingly, one patient (COVGEN-27) carried four rare heterozygous variants (two truncating and two missense) in *IRAK3, LY9, TLR3*, and *TRAF3*, while another patient (COVGEN-27) had three variants (two truncating and one missense) in *NLRP12, TLR6*, and *IRF3* (**Table 3**).

Overall, 19 patients with MIS-C (42.2%; 95% CI, 29.0%-56.7%) had rare truncating or missense variants (**Figure 4A**), and seven of those patients (37%; 95% CI, 19%-59%) had more than one variant (**Figure 3B**). Our combined enrichment analysis for both missense and truncating variants showed that patients with MIS-C have a significant burden of such variants relative to the control group.

**Figure 4.**
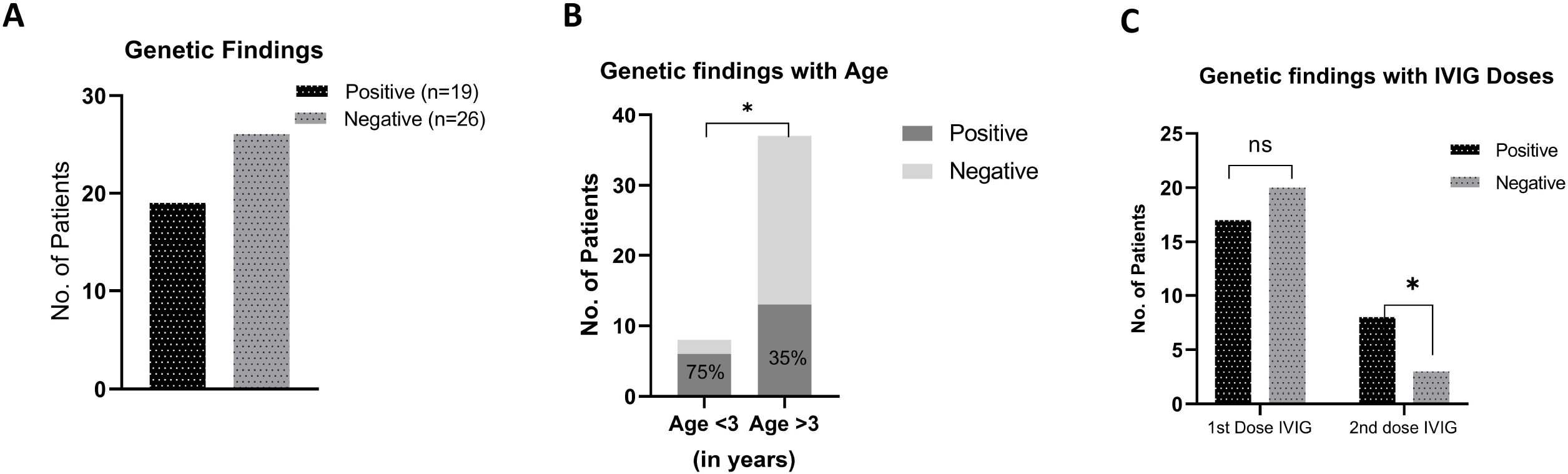
Genetic findings and associations with age and response to treatment. **A**, Number of patients with (black filled bar) or without (grey filled bar) genetic variants, y axis represents number of patients. **B**, Association of genetic findings with MIS-C patients’ age. Among MIS-C patients who were less than 3 years of age (total 8), the majority (75%) had a genetic finding (dark grey). On the other hand, 35% of patients older than 3 years had genetic variants, y axis represents number of patients, and x axis represent age in years. **C**, Association between genetic status and intravenous immunoglobulin (IVIG) treatment. While most patients received one dose of IVIG, a significantly higher proportion of patients with genetic findings (8/19, 42%) received a second dose compared to those without genetic variants (3/26, 11%). *P < 0.05 by Fisher’s Exact test.

### Pathway and Protein Network Analysis

Pathway analysis pinpointed the Toll-like receptor signaling and the interferon-mediated immune response pathways as the main pathways altered in MIS-C patients (**Table 3**). We obtained a protein-protein interaction (PPI) network with 21 nodes (genes) and 49 edges (functional associations) among these, *IFNB1, IFNA21, IFNA4, IFNA6, IFNAR2, IFI44, IFI44L*, and *TLR6* were among the highest connected nodes indicating the role of *IFN*-mediated immune responses (**Supplementary Figure 1**).

### Genetic associations with clinical and demographic characteristics of patients with MIS-C

We tested the association between the genetic status of patients with MIS-C and their demographic, clinical and laboratory findings (**Figure 1**). Patients who were less than 3 years of age were more likely to harbour (P<0.05, Fisher’s exact test) a rare variant in the immune-related genes (**Figure 4B**). Clinically, there was no significant association between genetic findings and symptoms (**Supplementary Table 3** and **4**). Most laboratory markers were similarly abnormal in all patients with MIS-C, though there was a slight trend for a significantly higher ferritin in those with a genetic finding (**Table 2**).

### Management, hospital course and outcomes

All patients recovered after an average length of stay of around six and a half days. Nineteen patients (42%) were admitted to the pediatric intensive care unit (PICU). Thirty-seven patients (82%) received at least one high dose (2 gram/kg) of IVIG over 12 hours. Of the eleven patients who received two doses of the IVIG, eight had genetic findings (8/19 versus 3/26, P = 0.037, two-tailed Fisher’s exact test) (**Figure 4C** and **Table 4**) suggesting a possible resistance to treatment. Thirty-one patients were given corticosteroids, while thirty-six were given aspirin. Enoxaparin was received by 9 patients. There was no association between genetic status and treatment with corticosteroid, aspirin, enoxaparin, or oxygen intervention. Notably, of the seven patients who had more than one genetic variant, five were admitted to PICU and received intensive care treatment (two-invasive, three-noninvasive ventilatory support), four of them had cardiac dysfunction, and all of them had GI symptoms. There were no deaths amongst patients recruited in this study.

**Table 4:** Treatment, management, and outcomes.

|  | Positive genetic<br>Variation (N = 19) |  | Negative genetic<br>Variation (N = 26) |  | P-value | All (% of total 45 patients) |
| --- | --- | --- | --- | --- | --- | --- |
| Admission to PICU, N (% within group) | 9 (47.3%) |  | 10 (38.4%) |  | 0.7 | 19 (42.2%) |
| Oxygen Invasive, N (% within group) | 4 (21%) |  | 5 (26%) |  | >0.99 | 9 (20%) |
| Oxygen Non-Invasive, N (% within group) | 5 (26%) |  | 4 (15%) |  | 0.4 | 9 (20%) |
| First dose IVIG, N (% within group) | 17 (89%) |  | 20 (77%) |  | 0.4 | 37 (82%) |
| Second dose IVIG, N (% within group) | 8 (42%) |  | 3 (5.7%) |  | 0.03 | 11 (24%) |
| Aspirin, N (% within group) | 15 (79%) |  | 21 (80%) |  | 0.9 | 36 (80%) |
| Corticosteroid, N (% within group) | 15 (79%) |  | 16 (61%) |  | 0.3 | 31 (69%) |
| Enoxaprain, N (% within group) | 4 (21%) |  | 5 (19%) |  | 0.9 | 9 (20%) |
| Deceased, N (% within group) | 0% |  | 0% |  | - | % |
|  | Mean | S. D | Mean | S. D |  |  |
| Duration of the Stay | 7 | 4.39 | 6.19 | 5.26 | 0.38 | 44 (98%) |

Abbreviations: PICU, Pediatric Intensive Care Unit. IVIG, Intravenous immune globulin.

## Discussion

In this study we characterize the genomic landscape, phenotypic features, inflammatory and cellular markers, and clinical management and outcomes of the largest prospective cohort of patients with MIS-C from the Middle East. To our knowledge, this study is also the largest MIS-C cohort involving whole exome sequencing data from 45 patients, along with 25 SARS-COV-2 positive control patients who did not meet the criteria for MIS-C diagnosis.

Our cohort included patients who were mostly from the Middle East (66.7%) including 26 Arab descendants (57.7%), and 9 (20%) Asians. Patients of such backgrounds have long been underrepresented in genetic studies emphasizing the importance of our study in characterizing the genetic landscape of MIS-C disease in this cohort. Although clinical presentations and laboratory markers in this cohort were consistent with recently described MIS-C cohorts elsewhere, our analysis revealed significant enrichment of rare, likely deleterious variants mainly affecting the Toll-like receptor signaling and interferon-mediated immune response pathways in patients relative to age- and ethnically-matched controls and to the general population of diverse origins (including our internal cohort of healthy individuals). Additionally, seven out of the 19 patients (37%) had more than one variant, including two patients with four and three variants, respectively, further highlighting the significant burden of genetic variation in this cohort. Despite the relatively high consanguineous marriages in this part of the world, our analysis did not identify any rare homozygous variants which might predispose to MIS-C. It is possible that such variants might lead to severe immune-related disorders irrespective of SARS-CoV2 infection. Patients with such genetic variants or disorders would have been excluded in our study as almost all our patients did not have significant medical history before viral infection.

Our analysis showed that genetic findings were significantly associated with earlier onset of disease (<3 years), and with possible resistance to IVIG treatment in MIS-C patients. With one exception (fibrinogen), there were no statistically significant associations between inflammatory markers or clinical symptoms and genetic findings. Such associations cannot be ruled out for several reasons. First, our sample size might not be empowered to detect such associations. Second, our study was designed to mainly capture rare, relatively large effect variants in selected genes (N = 186). Therefore, genetic contribution due to deleterious variants outside the 186 studied genes or due to polygenic, small effect variants across the genome cannot be ruled out. However, larger sample sizes are still needed to capture such small effect contributions.

One limitation of this study is the lack of functional analyses to characterize the mechanism(s) through which the mutated genes contribute to MIS-C disease onset and/or progression. However, as shown in **Table 3**, most variants are highly concentrated to genes in the interferon (IFN), and toll-like receptor (TLR) pathways suggesting that disturbance of such pathways might underlie the cytokine storms and dysregulated inflammatory markers (**Table 2**) in those patients. Protein-protein network analysis (**Supplementary Figure 1**) further confirms the significant interaction and convergence of most mutated genes in this study. Although our genetic analysis revealed some overlap with the type I IFN pathway shown to be altered in patients with severe COVID-19 (18), the majority of identified variants, especially truncating ones, in MIS-C patients affect genes which do not overlap with those in patients with severe COVID-19. These results suggest that MIS-C and severe COVID-19 have distinct genetic determinants and molecular etiologies.

In summary, our study on patients from the Middle East shows that MIS-C has a genetic component and paves the way for additional studies designed to include larger numbers of patients from diverse backgrounds along with functional analyses to fully characterize the genetic contribution to this new disease entity.

## Data Availability

All data produced in the present study are available upon reasonable request to the authors

## Acknowledgements

This study was fully funded by Al Jalila Foundation, Dubai, United Arab Emirates (Grant # AJF202059.

## Figure Legends

**Supplementary Figure 1**. Protein-protein interaction (PPI) network representation of the genes with rare variants in MIS-C patients. Nodes represent genes, and edges represent functional or predicted association between strongly connected genes.

