## Supplementary figures and images for "Genomic and phenotypic characterization of Multisystem Inflammatory Syndrome in Children (MIS-C): a prospective multicenter study from the Middle East"

### Supplementary Figure 1

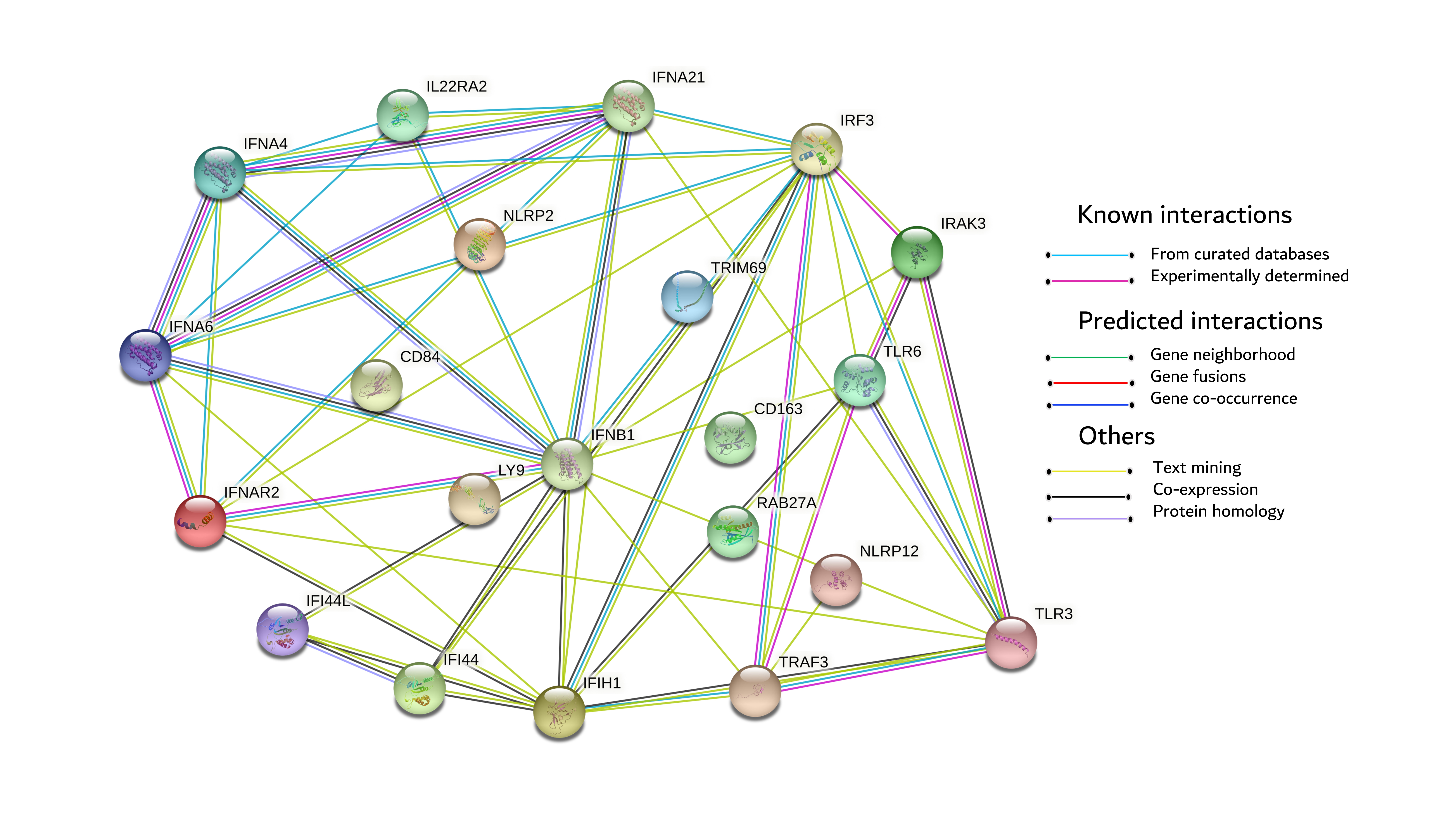

### Supplementary Table 1

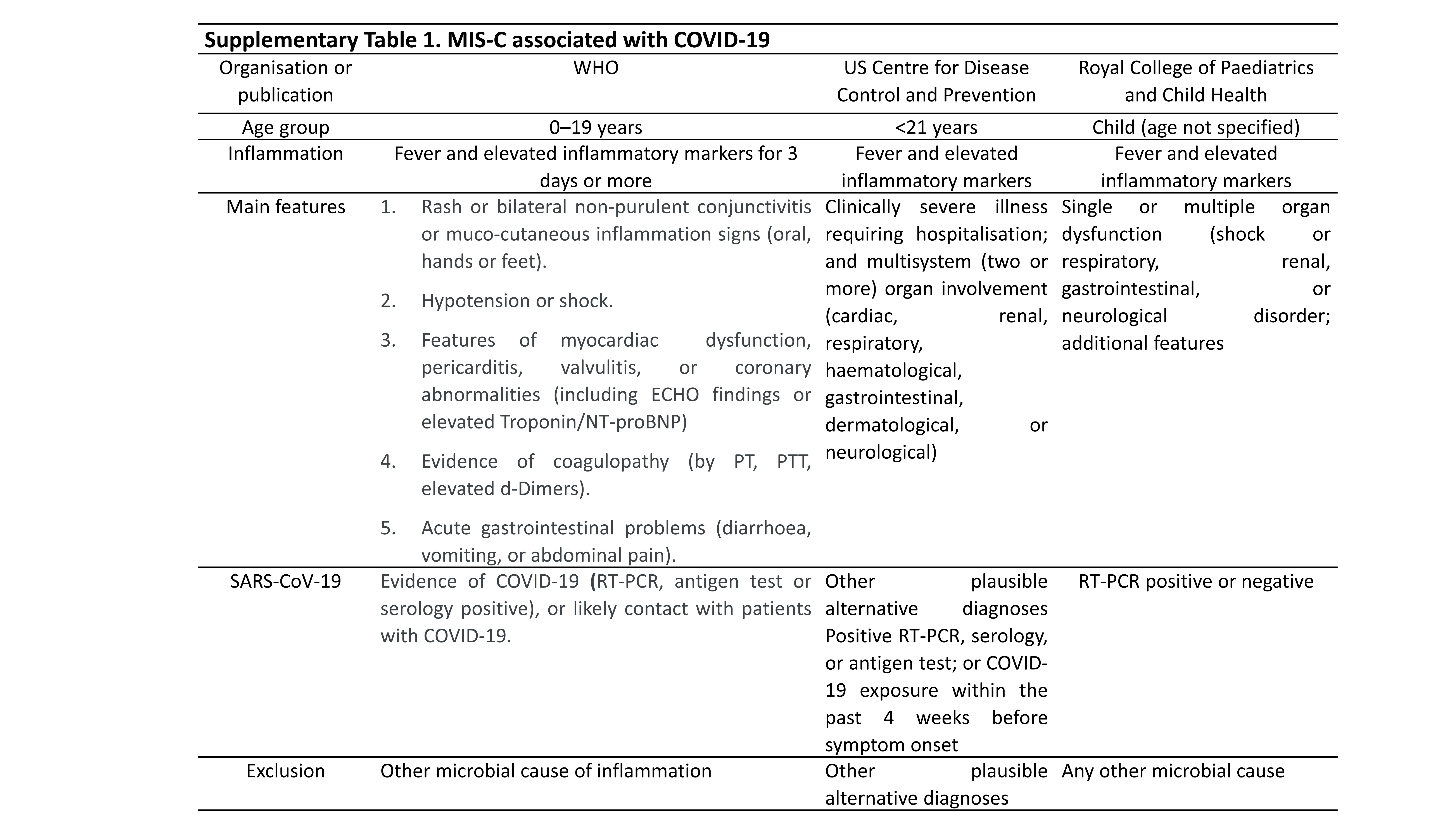

### Supplementary Table 2

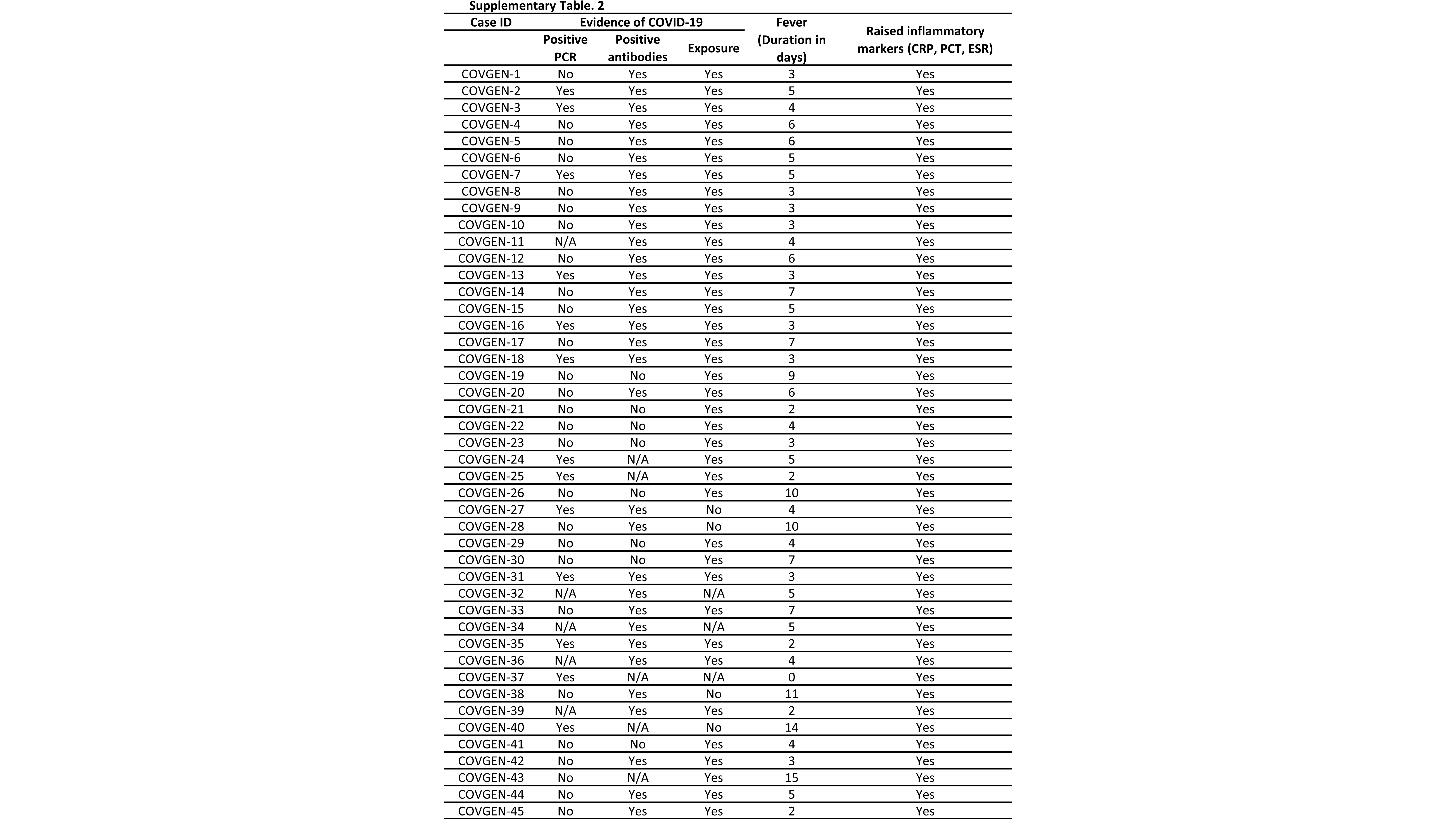

### Supplementary Table 3

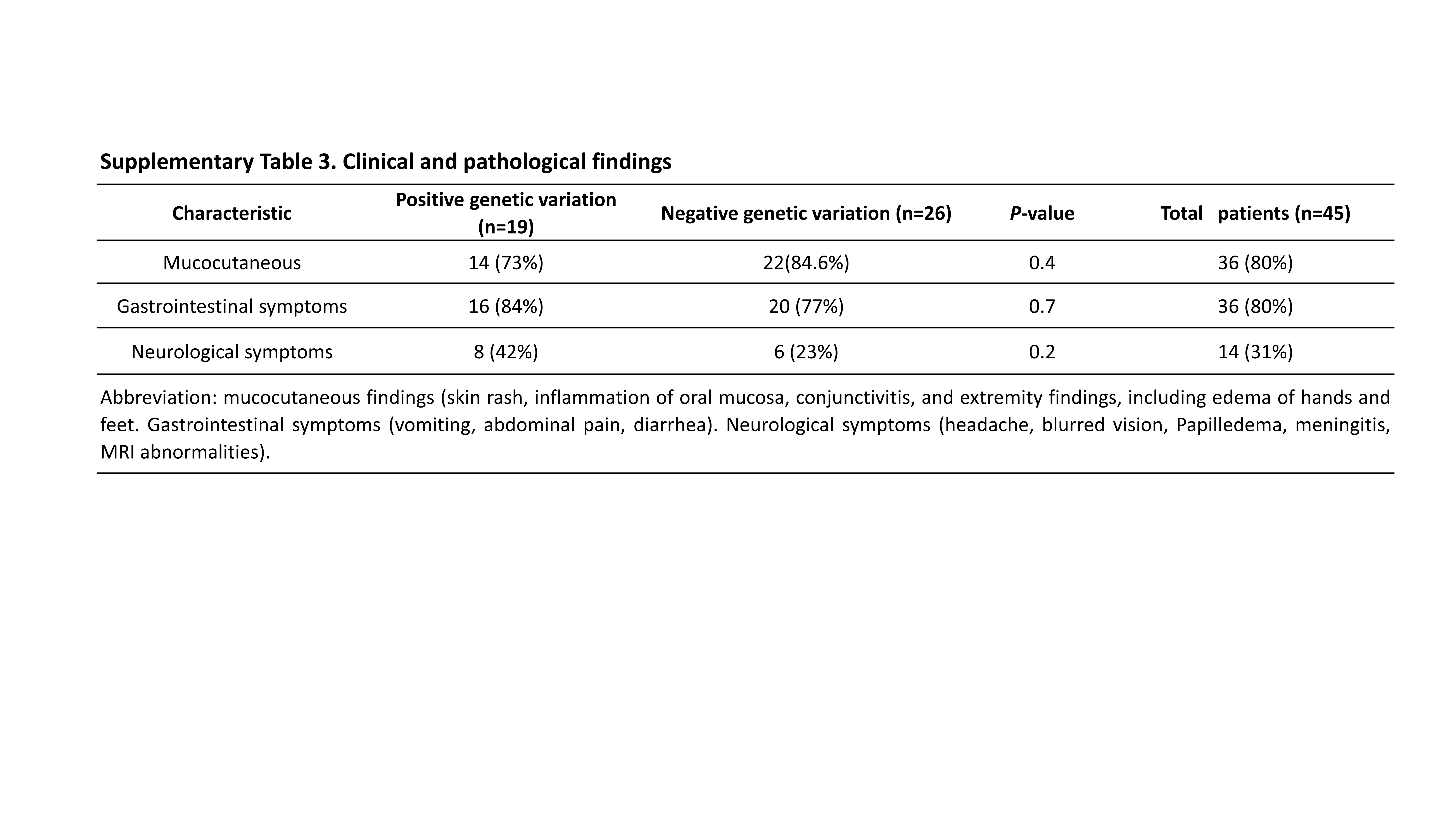
